## Supplement for "Optimal algorithms for controlling infectious diseases in real time using noisy infection data"

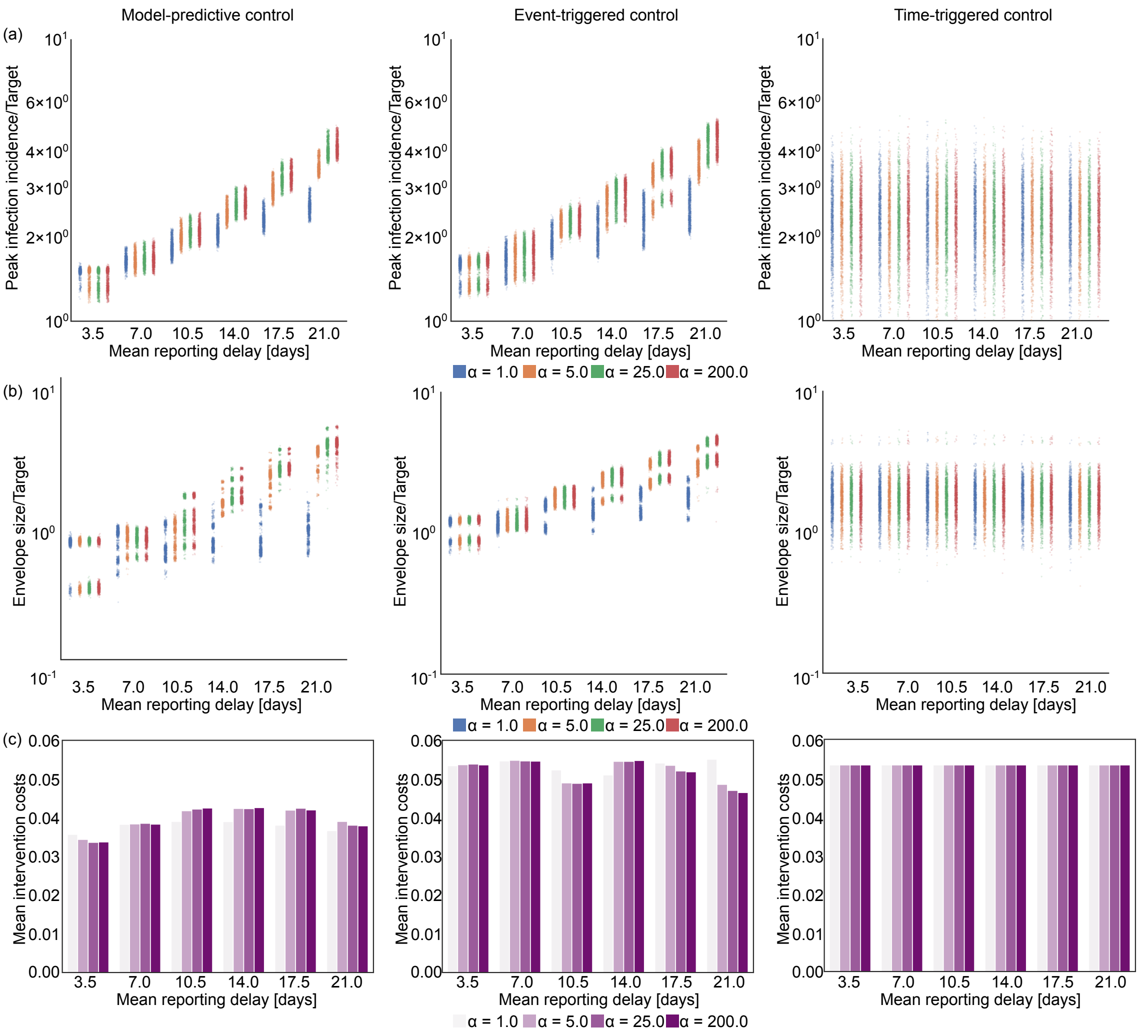


Figure 8: The impact of increasing reporting delay on optimal control. The left panels show results for our MPC algorithm, while the middle and right panels respectively present equivalent outputs under event-triggered (lockdown and relaxation thresholds of $C_{\mathrm{LD}} = 3000$ new cases/day, and $C_{\mathrm{relax}} = 3500$ new cases/day) and time-triggered (cycle of 36 days lockdown, 21 days of no restrictions, starting on day 143) strategies. Row (a) shows scatterplots for peak incidence and row (b) illustrates the steady-state envelope size relative to the target incidence across different time delay distributions displayed in the top row of Fig. 10. The horizontal axis represents different mean reporting delays with colours depicting different dispersion levels. Larger values of the parameter $\alpha$ indicate more deterministic delays. Row (c) shows the mean intervention costs for each ensemble of 1000 epidemics, simulated under estimated parameters from Ebola virus disease.

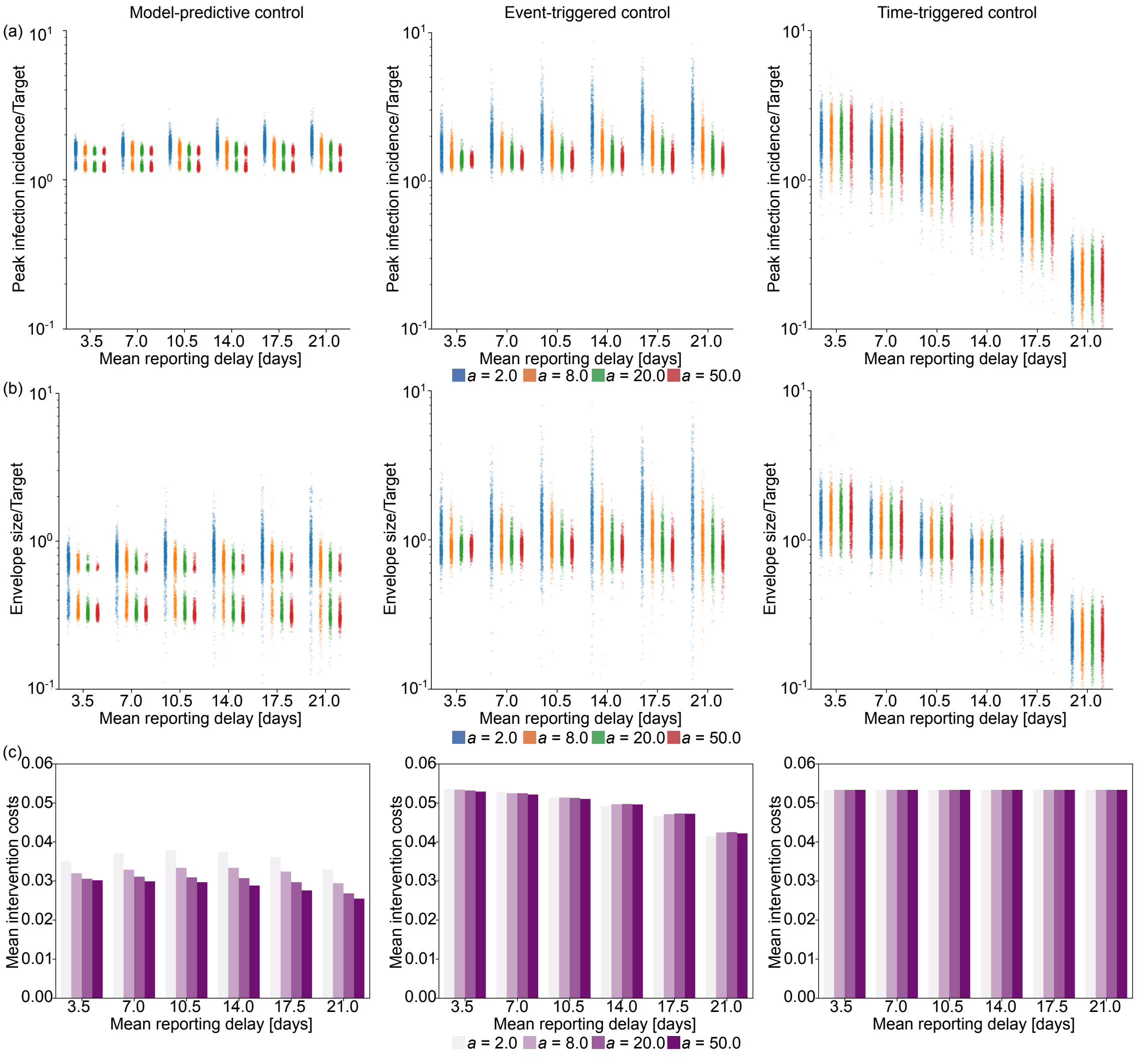


Figure 9: The impact of increasing under-reporting on optimal control. The left panels show results for our MPC algorithm, while the middle and right panels respectively present equivalent outputs under event-triggered (lockdown and relaxation thresholds of $C_{\mathrm{LD}} = 3000$ new cases/day, and $C_{\mathrm{relax}} = 3500$ new cases/day) and time-triggered (cycle of 36 days lockdown, 21 days of no restrictions, starting on day 143) strategies. Row (a) shows scatterplots for peak incidence, row (b) illustrates the steady-state envelope size relative to the target incidence for different reporting rate distributions displayed in the bottom row of Fig. 10. The horizontal axis represents different mean reporting ratios with colours depicting different dispersion levels. Larger values of the parameter $a$ belong to more deterministic case reporting distributions (i.e., constant reporting). Row (c) shows the mean intervention costs for each ensemble of 1000 epidemics, simulated under estimated parameters from Ebola virus disease.

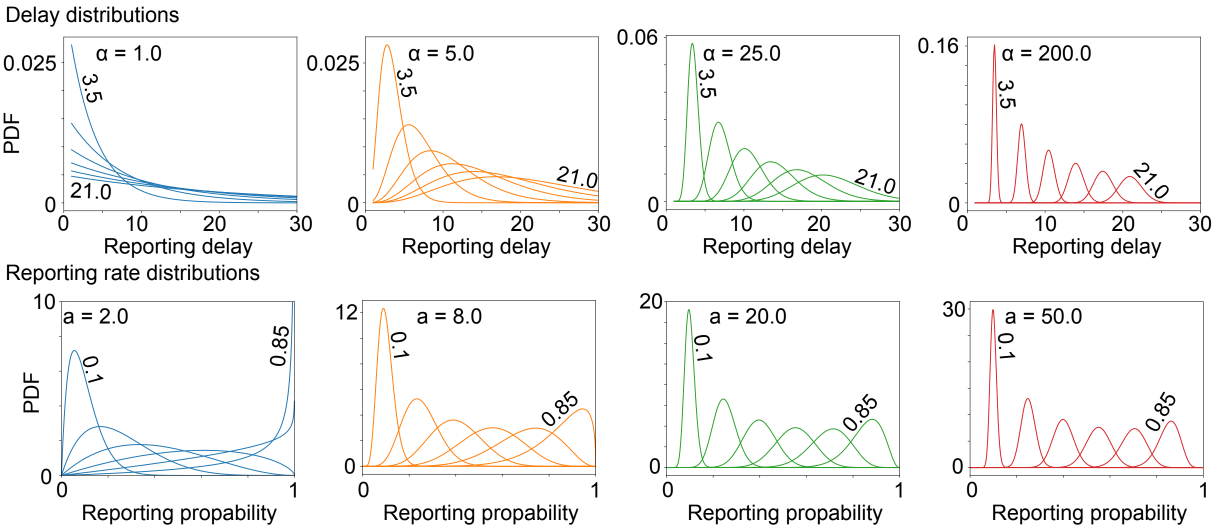


Figure 10: Probability density functions (PDFs) of the simulated surveillance noise. Top row: case reporting delay distributions modelled using Gamma distributions. The shape parameter $\alpha$ controls dispersion (larger $\alpha$ results in lower variance). Numbers above the curves indicate the mean of each distribution. Bottom row: infection reporting rate distributions modelled using Beta distributions. The parameter $a$ controls dispersion (larger $a$ results in lower variance). Numbers above the curves indicate the mean reporting rate.
